# Effects of Acute Normovolemic Hemodilution on Post-Cardiopulmonary Bypass Coagulation Tests and Allogeneic Blood Transfusion in Thoracic Aortic Repair Surgery: An Observational Cohort Study

**DOI:** 10.1101/2021.06.01.21258155

**Authors:** Domagoj Mladinov, Kyle W. Eudailey, Luz A. Padilla, Joseph B. Norman, Benjamin Leahy, Jacob Enslin, Keli Parker, Katherine F. Cornelius, James E. Davies

## Abstract

**Background and Aim:** Perioperative blood transfusion is associated with increased morbidity and mortality. Acute normovolemic hemodilution (ANH) is a blood conservation strategy associated with variable success, and rarely studied in more complex cardiac procedures. The study aim was to evaluate whether acute ANH improves coagulopathy and reduces blood transfusions in thoracic aortic surgeries.

**Methods:** Single-center observational cohort study comparing ANH and standard institutional practice in patients who underwent thoracic aortic repair from 2019 to 2021.

**Results:** 89 patients underwent ANH and 116 standard practice. There were no significant differences between the groups in terms of demographic or major perioperative characteristics. In the ANH group coagulation tests before and after transfusion of autologous blood showed decreased INR and increased platelets, fibrinogen, all with p<0.0005. Coagulation results in the ANH and control groups were not statistically different. The average number of transfused allogeneic products per patient was lower in the ANH vs control group: FFP 1.1 ±1.6 vs 1.9 ±2.3 (p=0.003), platelets 0.6 ±0.8 vs 1.2 ±1.3 (p=0.0008), and cryoprecipitate 0.3 ±0.7 vs 0.7 ±1.1 (p=0.008). Reduction in RBC transfusion was not statistically significant. The percentage of patients who received any transfusion was 53.9% in ANH and 59.5% in the control group (p=0.42). There was no significant difference in major adverse outcomes.

**Conclusions:** ANH is a safe blood conservation strategy for surgical repairs of the thoracic aorta. Laboratory data suggests that ANH can improve coagulopathy after separation from CPB, and significantly reduce the number of transfused FFP, platelets and cryoprecipitate.

## Introduction

Acute normovolemic hemodilution (ANH) is a blood conservation strategy that consists of intraoperative harvest of autologous whole blood (AWB) at the beginning of surgery, replacement of volume with crystalloid and/or colloid solutions, and transfusion of the autologous blood at the end of procedure. ^1 2^ Possible beneficial effects of hemodilution include preservation of red blood cell (RBC) and platelet mass during bleeding events, decreased cell damage and dysfunction by avoiding exposure to cardiopulmonary bypass (CPB), reduced negative impact on coagulation, and improved perfusion during CPB.^3 4 5^ The overwhelming majority of studies addressing ANH in cardiac surgeries include patients undergoing coronary artery bypass and/or valve surgery.^6 7^ However, thoracic aortic surgeries are associated with coagulopathy and high transfusion rates, therefore these particular type of surgeries may greatly benefit from ANH. A recent study in patients who underwent aortic repair with hypothermic circulatory arrest demonstrated significantly decreased blood product utilization.^8^ As of now, ANH in aortic surgeries remains relatively under studied, and its utilization is largely surgical team- or institution-dependent. Further, despite coagulation tests playing a significant role in guiding blood product transfusion, little is known about the effects of hemodilution on coagulation. Interestingly studies performed in cardiac surgeries other than aortic repairs suggest no clinically significant improvement in coagulation results.^9^ In this study we evaluated whether ANH improved coagulation and/or reduced allogeneic blood transfusion in surgical repairs of thoracic aortic aneurysms or dissections. We also assessed safety of this technique by analyzing multiple outcome measures.

## Methods

### Study Design and Patients

This observational cohort study was approved by our Institutional Review Board. The study included surgeries performed between 01/01/2019 and 01/15/2021, during which surgical and perfusion techniques, and transfusion practices remained unchanged. We identified all adult patients who underwent surgical repair of the thoracic aorta (with or without other concomitant procedures), including repeat sternotomies and emergent cases. The following exclusion criteria were applied: preoperative hematocrit < 30%, known pre-existing coagulopathy or ongoing anticoagulant or antiplatelet other than acetylsalicylic acid (ASA) use, left ventricular ejection fraction <35%, hemodynamic instability (requiring medication and/or mechanical support), use of extracorporeal membrane oxygenation (ECMO) support after surgery. Medical history, laboratory and procedural data were obtained by manual extraction from patient electronic medical records, and outcome data were obtained from the Society of Thoracic Surgeons National Database.

### Blood Management Practices

During the study period standard institutional blood conservation strategies included restrictive intraoperative fluid therapy, retrograde autologous priming of the CPB circuit (that was initially primed with 1.5 L of balanced crystalloid solution and 12.5 g of mannitol), ultrafiltration during CPB, cell salvage, and use of antifibrinolytics.

Whole blood removal was performed via a central venous catheter port by gravity drainage, immediately after the catheter placement. Blood was drained into citrate-phosphate-dextrose bags, and its weight measured on an electronic scale, with a maximum allowable weight of 450 grams per bag. The amount of removed blood was based on patient’s ideal body weight and starting hematocrit.^10^ Standard hemodynamic monitors and transesophageal echocardiography (TEE) were used to guide fluid replacement (crystalloid and/or colloid solution, at 1:2 ratio or less) and if needed the use of vasopressor medications. Autologous blood was kept in the operating room at room temperature, and transfused back after protamine administration.

In our intraoperative practice, typically the transfusion trigger for RBC includes hemoglobin < 7 g/dL. If during CPB transfusion was deemed necessary, a unit of allogeneic RBC was added to the CPB reservoir. Transfusion of other blood products was guided by standard coagulation laboratory tests and/or thromboelastography.

### Outcomes

The primary outcomes of the study were intraoperative transfusion of allogeneic blood products and changes in coagulation laboratory values after CPB. Secondary outcomes were postoperative transfusions during the index hospitalization, chest tube output during the first 12 hours after surgery, reoperation for bleeding, acute kidney injury, need for renal replacement therapy, ischemic stroke, duration of intubation, intensive care unit stay.

### Statistical Analysis

ANH and control patient demographics, peri-operative, clinical outcomes and laboratory variables were summarized (frequencies, percentages, medians, interquartile ranges [Q1-Q3], means and, standard deviations) and compared using chi-square and Fisher’s exact for categorical variables, student t-test and Wilcoxon’s rank sum test for continuous variables where appropriate. Paired t-test was used to compare the laboratory values after separation from CPB among ANH patients only. Analysis of variance was used to compare the differences in mean peri-operative hospital laboratory results in both ANH and controls. Statistical Analytical Software (SAS) 9.4 (SAS institute, Cary, NC) was used for the analysis and all statistical tests of a two-sided *P* value of < 0.05 were considered significant.

## Results

A total of 474 patients underwent thoracic aortic repair between January 2019 and January 2021. Inclusion criteria were met in 205 (43.2%) cases, of which 116 (56.6%) were treated according to standard institutional practices, and 89 (43.4%) were subjected to ANH in addition to the standard practices. Procedures included elective and emergent thoracic aortic repairs, and may have also included concomitant procedures such as valve repair/replacement and coronary artery bypass grafting. Demographic, historical, laboratory and procedural characteristics were not statistically different between the two groups, except for the proportion of emergent surgeries which was higher in the ANH compared to control group (17.9% vs 3.4% respectively, Table 1). The average volume of collected autologous blood was 960 ± 246 ml.

**Table 1.** Demographics, Pre- and Intra- operative characteristics for patients who received Acute Normovolemic Hemodilution (ANH) or Standard Institutional Practice without ANH

|  | ANH<br>N=89 (43.4%) | No ANH<br>N=116 (56.6%) | p value |
| --- | --- | --- | --- |
| Age (years) | 58.4 ± 13.4 | 57.8 ± 14.8 | 0.80 |
| Male (%) | 69 (77.5) | 82 (71.3) | 0.31 |
| Weight (kg) | 94.8 ± 22.9 | 92.2 ± 22.2 | 0.48 |
| BMI (kg/m<sup>2</sup>) | 30.1 ± 7.1 | 29.8 ± 6.3 | 0.82 |
| Past medical history |  |  |  |
| CAD/MI | 30 (33.7) | 36 (31.0) | 0.68 |
| HTN | 75 (84.3) | 91 (78.4) | 0.29 |
| CVA | 11 (12.4) | 13 (11.2) | 0.79 |
| COPD | 9 (10.1) | 22 (18.9) | 0.07 |
| DM | 11 (12.4) | 7 (6.0) | 0.11 |
| CKD | 13 (14.6) | 10 (8.6) | 0.17 |
| Current tobacco use | 21 (23.6) | 16 (13.8) | 0.07 |
| EuroSCORE (%) | 2.8 [1.6-5.1] | 3.0 [1.6-6.4] | 0.45 |
| ***Pre-Operative laboratory values*** |  |  |  |
| Creatinine (mg/dL) | 1.0 [0.9-1.2] | 1.0 [0.9-1.1] | 0.08 |
| BUN (mg/dL) | 18.6 ± 6.1 | 17.1 ± 6.9 | 0.09 |
| GFR < 60 mL/min | 22 (25.3) | 23 (19.8) | 0.35 |
| Hematocrit (%) | 41.1 ± 4.2 | 41.2 ± 4.8 | 0.78 |
| Hemoglobin (g/dL) | 13.8 ± 1.5 | 13.7 ± 1.7 | 0.56 |
| Platelet (10<sup>3</sup>/cmm) | 219.3 ± 64.4 | 203.9 ± 69.2 | 0.10 |
| INR | 1.03 [0.98-1.10] | 1.03 [0.99-1.08] | 0.90 |
| PT (seconds) | 13.7 ± 1.1 | 13.7 ± 1.1 | 0.89 |
| PTT (seconds) | 30.8 ± 8.1 | 33.6 ± 17.6 | 0.14 |
| ***Intra-Operative characteristics*** |  |  |  |
| Re-do sternotomy | 23 (25.8) | 28 (24.1) | 0.77 |
| Emergent | 16 (17.9) | 4 (3.4) | 0.0005 |
| Concomitant procedure<sup>^</sup> |  |  |  |
| CABG | 24 (26.9) | 22 (18.9) | 0.17 |
| AVR | 52 (58.4) | 74 (63.7) | 0.43 |
| MVR/TVR/PVR | 3 (3.1) | 6 (5.1) | 0.81 |
| Procedure time (min) | 260.4 ± 109 | 251.2 ± 124.0 | 0.57 |
| Cardiopulmonary bypass time (min) | 106.6 ± 54.3 | 98.9 ± 57.8 | 0.33 |
| Cross clamp time (min) | 71.8 ± 38.4 | 67.8 ± 37.7 | 0.46 |
| Circulatory arrest (yes) | 25 (28.1) | 26 (25.7) | 0.71 |
| Circulatory arrest time (min) | 23.5 ± 9.7 | 27.1 ± 18.5 | 0.38 |
| Cooling temp (Celsius) | 26.3 ± 2.4 | 26.3 ± 3.6 | 0.98 |

<sup>^</sup>All patients had aortic dissection and/or aortic aneurysm repair. In addition some patients had CABG and/or valve surgery Key: N (%); median [Q1-Q3]; mean ± SD; CAD=Coronary Artery Disease, MI=Myocardial Infarction, HTN=Hypertension, CVA=Cerebrovascular Accident, COPD=Chronic Obstructive Pulmonary Disease, DM=Diabetes Mellitus Type 2, CKD=Chronic Kidney Disease, BUN=Blood Urea Nitrogen, GFR=Glomerular Filtration Rate, INR=International Normalized Ratio, PT=Prothrombin time, PTT=Partial thromboplastin time, CABG=Coronary Artery Bypass Graft, AVR/MVR/TVR/PVR=Aortic/Mitral/Tricuspid/Pulmonary Valve Replacement/Repair

In the ANH group following separation from CPB and protamine administration, coagulation tests obtained before and after transfusion of autologous blood showed decreased INR from 1.84 [1.70-2.10] to 1.62 [1.49-1.76], p<0.0001, increased platelets from 122.9 ±48.8 to 140.1 ±44.8, 10^3/ccm, p<0.0001, and increased fibrinogen from 208.5 ±83.4 to 222.5 ±74.7 mg/dL, p=0.0002 (Table 2). Comparison of coagulation results in the ANH group after autologous transfusion and the control group after separation of CPB demonstrated a lower INR (1.62 [1.49-1.76] vs 1.74 [1.56-1.91], p=0.002), and a non-statistically significant difference in platelet count (140.1 ± 44.8 vs 129.8 ± 46.6, 10^3/ccm, p=0.12) and fibrinogen (222.5 ±74.7 vs 229.68 ± 57.7 mg/dL, p=0.46) (Table 2).

**Table 2.** Laboratory values after separation from cardiopulmonary bypass (CPB) in patients who received Acute Normovolemic Hemodilution (ANH) or Standard Institutional Practice without ANH

|  | ANH<br>N=89 (43.4%) |  |  | No ANH<br>N=116 (56.6%) |  |
| --- | --- | --- | --- | --- | --- |
|  | Pre ANH | Post ANH | *p* value* | Post CPB | *p* value** |
| Hematocrit (%) | 28.1 ± 4.1 | 30.4 ± 4.4 | <.0001 | 31.2 ± 5.1 | 0.23 |
| Hemoglobin (g/dL) | 9.1 ± 1.3 | 9.8 ± 1.6 | <.0001 | 10.1 ± 1.6 | 0.24 |
| Platelets (10<sup>3</sup>/cmm) | 122.9 ± 48.8 | 140.1 ± 44.8 | <.0001 | 129.8 ± 46.6 | 0.12 |
| Fibrinogen (mg/dL) | 208.5 ± 83.4 | 222.5 ± 74.7 | 0.0002 | 229.68 ± 57.7 | 0.46 |
| INR | 1.84 [1.70-2.10] | 1.62 [1.49-1.76] | <.0001 | 1.74 [1.56-1.91] | 0.002 |
| PT (seconds) | 21.7 ± 3.2 | 19.3 ± 4.9 | <.0001 | 19.8 ± 2.3 | 0.38 |
| PTT (seconds) | 37.0 ± 11.4 | 39.3 ± 15.5 | 0.07 | 39.0 ± 19.5 | 0.89 |

Key: N (%); medians [Q1-Q3]; mean ± SD, INR=International Normalized Ratio, PT=Prothrombin time, PTT=Partial thromboplastin time
\*P value for Pre ANH and Post ANH comparison among those who received ANH (n=89)
\*\*P value comparing the laboratory values Post ANH among those who received ANH vs the Post CPB values among those who received standard institutional practice without ANH

The average number of intraoperative allogeneic blood product administration per patient in the ANH vs control group was: PRBC 0.49 ±1.1 vs 0.62 ±1.4 (p=0.44), FFP 1.1 ±1.6 vs 1.9 ±2.3 (p=0.003), platelets 0.6 ±0.8 vs 1.2 ±1.3 (p=0.0008), and cryoprecipitate 0.3 ±0.7 vs 0.7 ±1.1 (p=0.008). The percentage of patients who received any allogeneic transfusion was 53.9% in ANH and 59.5% in the control group (p=0.42), and the break down by blood components showed statistically lower transfusion rate for cryoprecipitate only (Table 3). In the postoperative period through the index hospitalization, the average number of allogeneic blood transfusion in the ANH vs control group was: PRBC 0.5 ±1.0 vs 0.9 ±2.0 (p=0.04), FFP 0.3 ±0.9 vs 0.5 ±1.5 (p=0.17), platelets 0.2 ±0.7 vs 0.3 ±0.7 (p=0.51), and cryoprecipitate 0.1 ±0.5 vs 0.2 ±0.7 (p=0.30). The proportion of patients who received any allogeneic transfusion postoperatively was 53.9% in ANH and 59.5% in the control group (p=0.42), with no statistical difference when individual blood components were analyzed (Table 3).

**Table 3.**
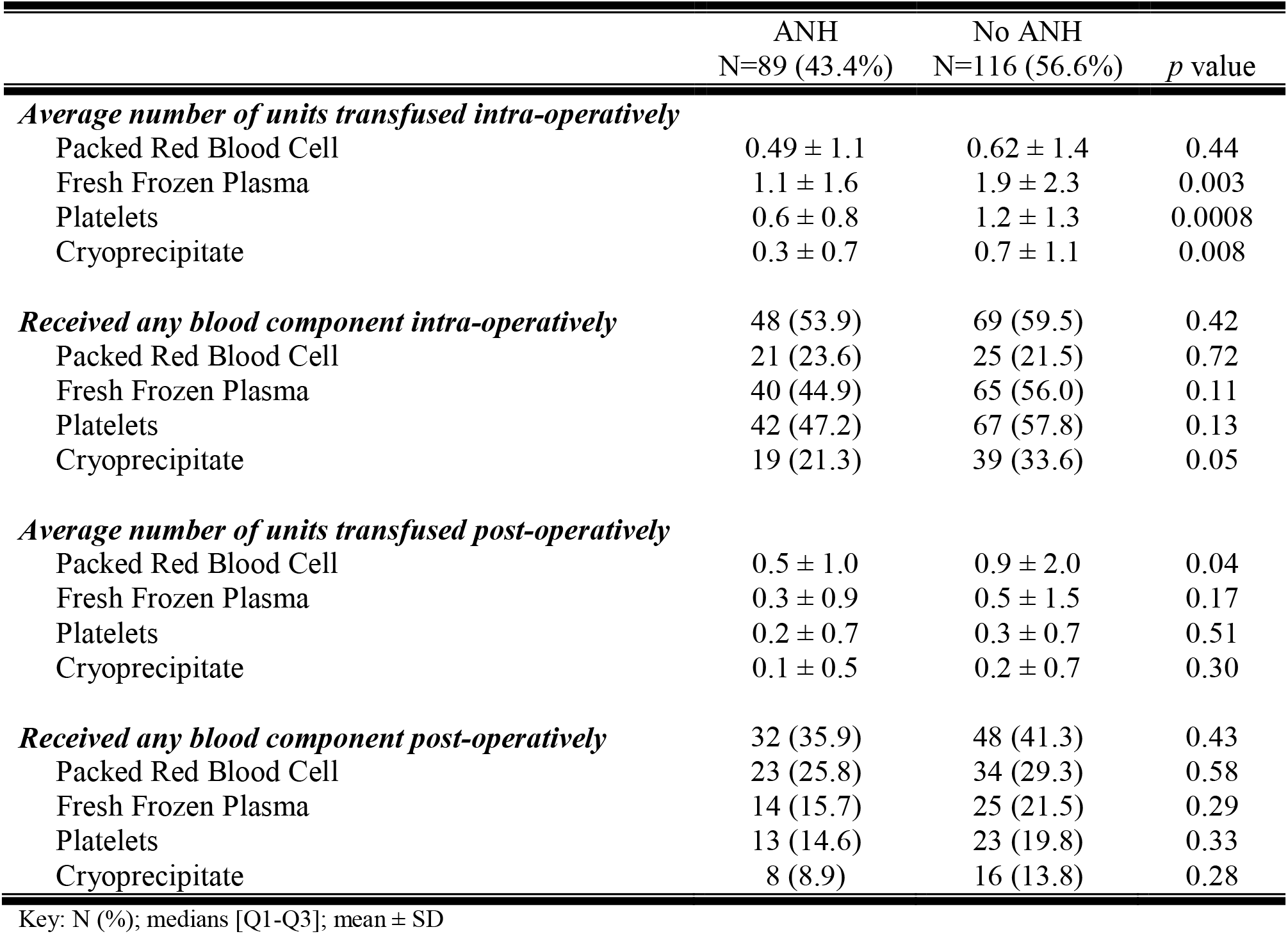
Transfusion rates and blood product utilization of cardiac surgery patients who received Acute Normovolemic Hemodilution (ANH) to Standard Institutional Practice without ANH

Differences in mean hemoglobin, platelets, INR and PTT values between the two groups at various perioperative time points are shown in Figure 1. Postoperative laboratory values were analyzed at various time points based on data availability: baseline, post ANH for those who underwent ANH or post CPB for those with standard institutional practice, immediately upon admission to the intensive care unit (ICU), 8-36 hours after admission, and postoperative day 7 or hospital discharge. The differences in mean laboratory values across the hospital continuum for both ANH and standard institutional practice were significant (all p=<.0001), except for PTT changes among those with standard institutional practice. Comparison between control vs ANH was statistically significant for hemoglobin (10.4 ± 1.5 vs 9.8 ± 1.8, g/dL, p=0.01) and hematocrit (31.8 ± 4.9 vs 30.1 ± 4.8, %, p=0.01) at the second time point; and for hemoglobin (9.9 ± 1.4 vs 9.5 ± 1.1, g/dL, p=0.01), creatinine (0.9 [0.8-1.1] vs 1.0 [0.8-1.2], mg/dL, p=0.05) and BUN (22.7 ± 9.7 vs 26.1 ± 13.5, mg/dL, p=0.05), at the last time point, respectively (Table 4).

**Table 4.** Post-operative laboratory values in patients who received Acute Normovolemic Hemodilution (ANH) or Standard Institutional Practice without ANH

|  | ANH<br>N=89 (43.4%) | No ANH<br>N=116 (56.6%) | *p* value |
| --- | --- | --- | --- |
| ***Intensive Care Unit (upon arrival)*** |  |  |  |
| Hematocrit (%) | 31.5 ± 4.5 | 32.3 ± 5.4 | 0.29 |
| Hemoglobin (g/dL) | 10.2 ± 1.5 | 10.5 ± 1.7 | 0.29 |
| Platelets (10<sup>3</sup>/cmm)* | 142.7 ± 37.9 | 148.2 ± 49.9 | 0.61 |
| INR* | 1.39 [1.32-1.49] | 1.34 [1.13-1.46] | 0.29 |
| PT (seconds)* | 16.6 ± 1.6 | 15.5 ± 2.7 | 0.09 |
| PTT (seconds)* | 38.1 ± 9.1 | 37.2 ± 5.3 | 0.65 |
| ***Intensive Care Unit (8-36 hrs)*** |  |  |  |
| Hematocrit (%) | 30.1 ± 4.8 | 31.8 ± 4.9 | 0.01 |
| Hemoglobin (g/dL) | 9.8 ± 1.8 | 10.4 ± 1.5 | 0.01 |
| Platelets (10<sup>3</sup>/cmm) | 128.2 ± 39.6 | 130.3 ± 45.2 | 0.73 |
| GFR < 60 mL/min | 26 (29.2) | 26 (22.6) | 0.28 |
| Creatinine (mg/dL) | 1.0 [0.8-1.3] | 0.9 [0.8-1.1] | 0.06 |
| BUN (mg/dL) | 20.3 ± 8.6 | 18.6 ± 7.6 | 0.15 |
| ***Post-operative day 7 or hospital discharge*** |  |  |  |
| Hematocrit (%) | 28.2 ± 3.3 | 29.1 ± 4.2 | 0.08 |
| Hemoglobin (g/dL) | 9.5 ± 1.1 | 9.9 ± 1.4 | 0.01 |
| Platelets (10<sup>3</sup>/cmm) | 210.5 ± 93.6 | 204.2 ± 76.0 | 0.60 |
| GFR < 60 mL/min | 23 (26.1) | 22 (18.9) | 0.22 |
| Creatinine (mg/dL) | 1.0 [0.8-1.2] | 0.9 [0.8-1.1] | 0.05 |
| BUN (mg/dL) | 26.1 ± 13.5 | 22.7 ± 9.7 | 0.05 |

Key: N (%); medians [Q1-Q3]; mean ± SD
\*60-75% of the data is missing
INR=International Normalized Ratio, PT=Prothrombin time, PTT=Partial thromboplastin time, GFR=Glomerular Filtration Rate, BUN=Blood Urea Nitrogen

**Figure 1.**
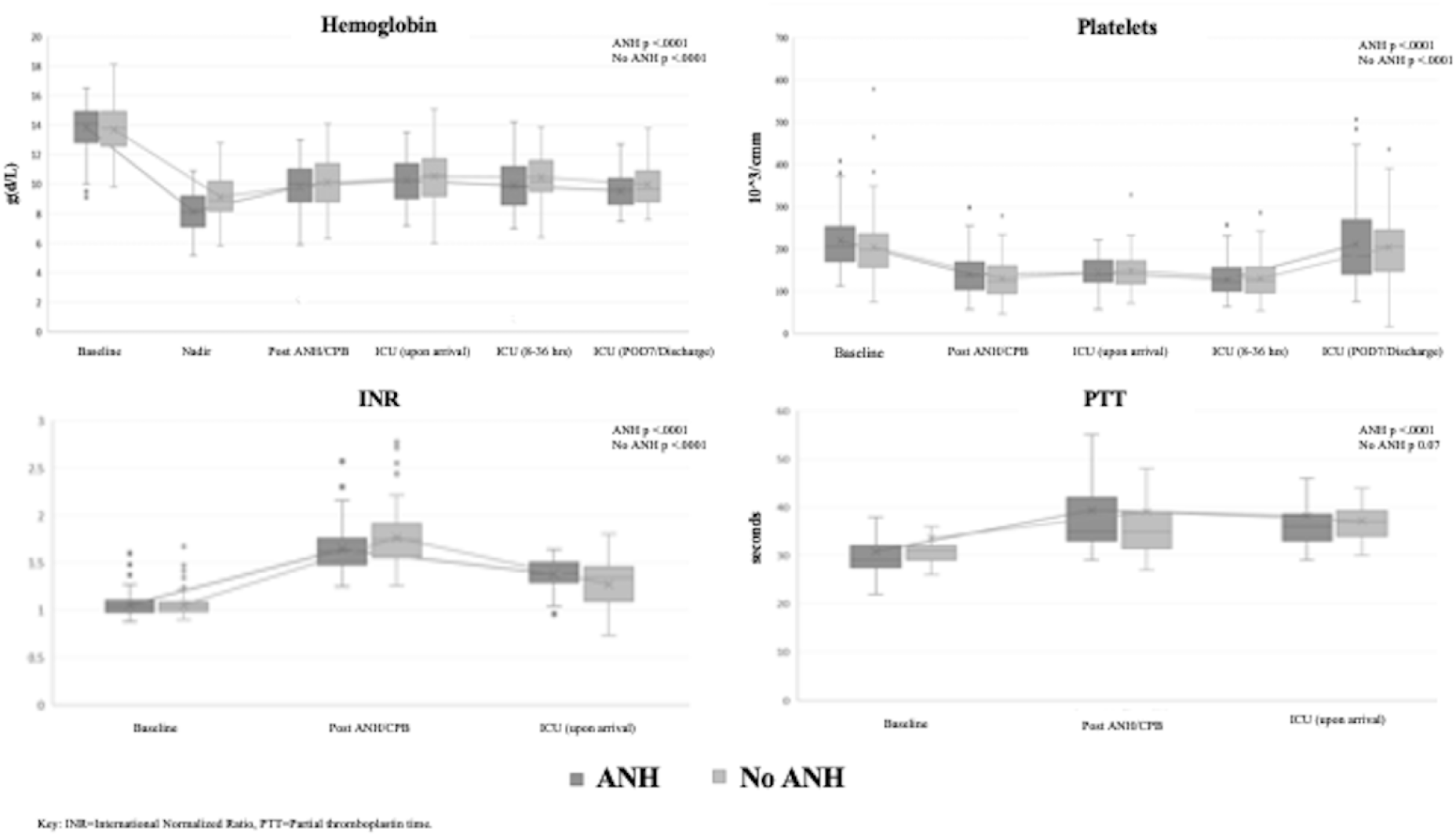
Differences in mean hemoglobin, platelets, INR and PTT values between the ANH and control groups at various perioperative time points: baseline, post-ANH (for the ANH group) or post-CPB (for the control group), immediately upon admission to the intensive care unit (ICU), 8-36 hours after admission, and postoperative day 7 or hospital discharge. All differences in mean laboratory values for both ANH and control were significant (p=<0.0001), except for PTT.

Secondary outcomes including post-operative 12h chest tube output, reoperation for bleeding, peak creatinine during hospitalization, need for renal replacement therapy, ischemic stroke, duration of intubation and intensive care unit length of stay were not significantly different between ANH and no ANH groups (Table 5).

**Table 5.** Post-operative surgical outcomes for patients who received Acute Normovolemic Hemodilution (ANH) or Standard Institutional Practice without ANH

|  | ANH<br>N=89 (43.4%) | No ANH<br>N=116 (56.6%) | *p* value |
| --- | --- | --- | --- |
| 12 hour chest tube output (ml) | 467.6 ± 250.1 | 498.7 ± 353.7 | 0.38 |
| Re-operation for bleeding | 0 (0) | 2 (1.7) | - |
| Mechanical ventilation (hours) | 5.0 [3.9-6.8] | 4.5 [3.3-7.1] | 0.25 |
| ICU length of stay (hours) | 31.9 [22.8-75.1] | 25.9 [21-48] | 0.15 |
| Peak creatinine (mg/dL) | 1.2 [1.0-1.6] | 1.1 [0.9-1.4] | 0.14 |
| Need for RRT | 0 (0) | 4 (3.4) | - |
| Ischemic Stroke | 5 (5.6) | 1 (0.9) | 0.08 |

Key: N (%); medians [Q1-Q3]; mean ± SD
ICU=Intensive Care Unit, RRT=Renal replacement therapy

## Discussion

Our results show that intraoperative hemodilution before procedure initiation, and return of the collected autologous blood after separation from CPB was associated with improved INR, platelet count and fibrinogen level. It was also associated with reduced intraoperative transfusion of allogeneic FFP, platelets and cryoprecipitate, while reduction in RBC utilization did not reach statistical significance. Postoperative reduction in blood transfusion was not demonstrated. Finally, ANH was not associated with significant differences in incidence of adverse events.

Blood product transfusion in cardiac surgery have been associated with increased morbidity and mortality, as well as healthcare cost. ^11 12 13^ Various blood conservation strategies are widely accepted and commonly used in cardiac surgery, such as intraoperative sell salvage, retrograde priming of CPB circuit, and antifibrinolytic therapy. The Society of Thoracic Surgeons recommends considering acute normovolemic hemodilution, but also acknowledges that its usefulness has not been well established – its use is supported by level of evidence B, with a recommendation class IIb. ^14^ Mixed results may be secondary to variability in case types, AWB volume and/or transfusion practices. ^15 16^ Most studies addressing the practice of ANH have focused on RBC transfusion rates rather than other blood components, and also elective valve repairs/replacements and coronary revascularizations rather than complex aortic repairs. Hesitation to performing hemodilution in more complex and higher risk cardiac surgeries is understandable, as hemodilution may be associated with hemodynamic instability and organ injury. ^17 18^ However, surgeries associated with significant coagulopathy and higher risk of bleeding may benefit most from this blood conservation strategy, as suggested by a recently published study by Geube et al. ^8^ Careful selection of patients and procedures may be crucial in successfully implementing ANH and clearly demonstrating its benefits.

In this study we selected only patients undergoing thoracic aortic dissection or aneurysm repairs, with or without other concomitant procedures, and also included emergent cases. Preoperative and intraoperative characteristic between the two groups were not statistically different, except for the proportion of emergent cases which was larger in the ANH group (17.9% in ANH, and 3.4% in controls, p=0.0005). As emergent procedures are associated with worse outcomes, this study suggests safety and benefits of hemodilution in high risk cardiac surgeries. ^19^

Benefits of ANH on decreased allogeneic blood transfusion was seen almost exclusively in the intraoperative period. This results are not surprising as return of AWB after separation of CPB is considered to improve intraoperative coagulopathy, which is supported by our intraoperative laboratory findings (Table 2). In the ANH group after separation from CPB, transfusion of AWB improved coagulopathy as assessed by standard coagulation tests. Comparison of those test between the ANH and control group did not demonstrate significant difference, however the control group received significantly more allogeneic blood products during this period, which likely contributed to the similar correction of laboratory values. Also, hemoglobin and coagulation test results were similar between the groups upon arrival in the ICU. Importantly, we found lack of significant difference in coagulation results between the two groups reassuring, as they suggested consistency in transfusion practice (transfusion triggers) across all patients. Difference in hematocrit and hemoglobin at 8-36h; and hemoglobin, creatinine and BUN at post-operative day 7/ hospital discharge, reached statistical significance, however the small difference in their absolute values is unlikely to be physiologically relevant. In this observational study transfusion practices remained unchanged and not protocolized. Although authors believe that point of care coagulation tests do have a role in assessing coagulopathy and improving transfusion practices, in order to avoid change in practice and potential introduction of bias, point of care tests were not mandated. ^20 21^

Primary outcomes show that utilization of ANH is associated with intraoperative reduction in FFP, platelets and cryoprecipitate, but not PRBC administration. This was somewhat surprising as other studies demonstrate also reduction in PRBC transfusions. ^8 15^ Such outcomes may be a result of our intraoperative ability to salvage red blood cell, while plasma coagulation factors and platelets are lost with surgical bleeding. This findings emphasize the value autologous blood may have in improving coagulopathy, which is supported by the coagulation studies discussed earlier. Another interesting finding was that ANH was not associated with a decreased proportion of patients who received PRBC, FFP or platelets. Overall, ANH was associated with a reduced amount of transfused allogeneic blood, but did not reduce the proportion of patients exposed to allogeneic blood intraoperatively.

Our study demonstrated that ANH did not affect chest tube output during the first 12h after surgery, nor the frequency of re-operation for bleeding, consistent with the study by Geube at al. ^8^ Adverse events or complications such as prolonged intubation, ICU stay, acute kidney injury (peak creatinine, need for renal replacement therapy) and stroke were not statistically different between the ANH and control groups. This results are consistent with other studies demonstrating safety of ANH in complex thoracic aortic repairs. ^8 22^

## Limitations

Limitations for observational studies apply, and despite stringent inclusion and exclusion criteria, selection bias remains plausible. The number of enrolled patients is limited as the study spanned over only two years, in an attempt to maintain the clinical practices both consistent and contemporary. We opted for a pragmatic approach, so transfusion practices were not protocolized, but remained unchanged throughout the study period. Also, according to institutional practices, standard coagulation tests rather than thromboelastometry were predominantly used. The limited number of patients did not allow for propensity score matching, however the ANH and control groups did not differ in any major demographic, medical history, laboratory or procedural characteristic, other than having a larger proportion of emergent cases in the ANH group (Table 1). Finally, the study is a single centered study that may lack generalizability to other centers and had a small sample that may have prevented us from finding meaningful differences of our secondary outcomes, such as re-operation for bleeding and need for renal replacement therapy.

## Conclusion

Our study suggests that intraoperative hemodilution before procedure initiation, and return of the collected autologous blood after separation from CPB may decrease intraoperative, but not postoperative transfusion of allogeneic blood products in surgical repairs of the thoracic aorta. Further, our results demonstrate safety of ANH during such high risk surgical procedures. The study also suggests safety of ANH in emergent cases, but is not powered enough to support that statement. Finally, laboratory results indicate that in thoracic aortic repairs, ANH improves coagulopathy after separation from CPB. Larger trials are needed in the future, to better evaluate effects of ANH on coagulopathy and blood transfusions in high risk cardiac surgical procedures.

## Data Availability

The data that support the findings of this study are available on request from the corresponding author.

## Abbreviations

ANH: acute normovolemic hemodilution
AWB: autologous whole blood
CPB: cardiopulmonary bypass
ECMO: extracorporeal membrane oxygenation
FFP: fresh frozen plasma
INR: international normalized ratio
PT: Prothrombin time
PTT: Partial thromboplastin time
RBC(s): red blood cell(s)
TEE: transesophageal echocardiography

## Notes

**<u>Conflict of Interests</u>**: The authors have no conflicts of interests to disclose.

**<u>Funding</u>**: No funding was received for the study.

### Competing Interest Statement

The authors have declared no competing interest.

### Funding Statement

No external funding was received for this study.

### Author Declarations

The study was approved by the University of Alabama at Birmingham Institutional Review Board (Project Number: IRB-300005436; Approval Date: 06/12/2019).

